# Association between antihypertensive combinations and postoperative mortality and functional decline: a nationwide survey of Japanese adults undergoing major surgeries

**DOI:** 10.1101/2024.03.14.24304265

**Authors:** Rena Suzukawa, Shintaro Mandai, Yuta Nakano, Shunsuke Inaba, Hisazumi Matsuki, Yutaro Mori, Fumiaki Ando, Takayasu Mori, Koichiro Susa, Soichiro Iimori, Shotaro Naito, Eisei Sohara, Tatemitsu Rai, Kiyohide Fushimi, Shinichi Uchida

## Abstract

**Background:** Considering the limited information available regarding the impact of antihypertensive classes on mortality and physical function during hospitalization, we aimed to clarify the impact of six antihypertensive classes, namely thiazide/thiazide-like diuretics (TH), calcium receptor blockers (CCBs), renin–angiotensin–aldosterone system inhibitors (RASis), mineral corticoid receptor antagonists, α-blockers, and β-blockers, on outcomes in adult patients undergoing major surgeries.

**Methods:** This study was a subanalysis of a nationwide observational cohort study involving Japanese adults undergoing major surgeries from 2018 to 2019 using an administrative claims database. We recruited 473,327 antihypertensive medication users and 376,583 nonusers aged ≥50 years who underwent six different types of surgeries, including coronary artery bypass grafting (CABG), thoracic lobectomy, orthopedic surgery, hepatopancreatobiliary surgery, gastrointestinal resection, and urological surgery. The risk for overall death or functional decline, defined as a ≥5-point decrease in the Barthel Index score during hospitalization, was determined using multivariable logistic regression models.

**Results:** All-cause inhospital deaths occurred in 5,777 (1.2%) users and 2,657 (0.7%) nonusers. Functional decline was observed in 42,930 (9.2%) users and 22,550 (6.0%) nonusers. Among single class users, RASi use had a multivariable odds ratio (OR) of 0.77 (95% confidence interval (CI) 0.63–0.93 vs. TH) for the composite of mortality and functional decline. β-Blocker use was associated with an increased risk for functional decline (OR 1.27, 95% CI 1.01–1.60 vs. TH). Among the recipients of the two medication classes, TH/RASi usage was associated with the lowest risk for composite outcome (OR 0.68, 95% CI 0.60–0.77 vs. TH/CCB). Among the recipients of the three or more medication classes, TH/CCB/RASi or TH/CCB/RASi/other displayed the lowest odds for composite outcome (OR 0.72, 95% CI 0.49–0.82 vs. TH/CCB/other; OR 0.63, 95% CI 0.49–0.82 vs. TH/CCB/others). A stratified analysis revealed that RASi users had a lower OR for the composite outcome after major surgery categories except CABG than non-RASi users.

**Conclusions:** RASis were associated with decreased risk of postoperative mortality and functional decline regardless of the number of antihypertensive classes or surgery type. Managing hypertension through multidrug combinations, including RASis, may mitigate mortality and loss of physical function during the perioperative period.

**Clinical Perspective:** What is new?

- This nationwide observational cohort study of Japanese adults undergoing major surgeries from 2018 to 2019 using an administrative claims database showed that all-cause inhospital deaths occurred in 5,777 (1.2%) antihypertensive users and 2,657 (0.7%) nonusers, whereas functional decline was observed in 42,930 (9.2%) antihypertensive users and 22,550 (6.0%) nonusers.
- We found that an increase in the number of antihypertensive classes used, indicative of patients with treatment-resistant hypertension, was associated with a higher risk of mortality and loss of physical function, partly attributed to loop diuretic use for congestion.

What are the clinical implications?

- This study determined combinations of antihypertensive drugs that potentially improve the outcomes of antihypertensive users undergoing major surgeries, with the favorable regimens including RASis independent of the number of antihypertensive classes used.
- After undergoing all major surgery categories except CABG, patients on RASis were at a lower risk of death and functional decline than those who were treated with other antihypertensive classes.

## Introduction

The proportion of patients with hypertension continues to increase globally. The number of adult patients with hypertension doubled from 1990 to 2019, affecting nearly 1,300 million people worldwide [1]. Although recent advances in antihypertensive medications and their widespread availability have improved hypertension control in this population, the prevalence of treatment-resistant hypertension requiring a combination of multiple antihypertensive classes has continued to increase [2–5]. When caring for patients with hypertension, the prevention of chronic systemic organ dysfunction, hospitalization, acute kidney injuries (AKI), and death caused by acute hypertension (AHT) is of considerable importance [6]. We previously reported an increase in absolute death and urgent dialysis due to AHT from 2010 to 2019 among hospitalized Japanese patients [6]. Given the lack of comprehensive investigations on the effective combination of antihypertensive classes and the mostly empiric evidence available [4], understanding the effects of various antihypertensive medications and their management approaches for both in- and outpatients has been of notable interest among physicians.

The decline in activities of daily living (ADLs) and frailty related to hospitalization have also been an enormous health concern for inpatients, given the exponential increase in the aging population globally. Previous studies have established that the use of angiotensin-converting enzyme inhibitors (ACEis) has long-term effects on muscle strength [7,8]. The use of angiotensin II receptor blockers (ARBs) promotes enhanced exercise capacity among patients with heart failure [9]. In contrast, our studies showed that patients taking loop diuretics were at an increased risk of developing sarcopenia [10,11], with subsequent studies reporting the same correlation among patients with heart failure and hepatopathy [12–14]. However, no study has focused on the effects of antihypertensive class or loop diuretics during hospitalization under acute care settings, i.e., in patients with severe diseases (e.g., cardiovascular diseases [CVDs], infectious diseases, or major surgeries). We evaluated the impact of continued empirical antihypertensive drug administration during hospitalization on postoperative outcomes, including mortality and physical function, after major surgeries.

The current study aimed to determine the most effective monotherapy or combination therapy among six antihypertensive classes (i.e., thiazide/thiazide-like diuretics [TH], calcium receptor blockers [CCBs], renin–angiotensin–aldosterone system inhibitors [RASis], mineral corticoid receptor antagonists [MRAs], α-blockers, and β-blockers) and loop diuretics, which helps lower the risk for postoperative mortality and functional decline among patients aged 50 and older who underwent major surgeries during hospitalization. We also aimed to investigate whether the type of major surgery (i.e., coronary artery bypass grafting [CABG], thoracic lobectomy, orthopedic surgery, hepatopancreatobiliary surgery, gastrointestinal resection, and urological surgery) altered the association between antihypertensive class and postoperative outcomes.

## Methods

### Study design and participants

This study was a *posthoc* subanalysis of a national survey on postoperative outcomes among Japanese adults undergoing major surgeries [15]. We used an administrative claims database, the Diagnosis Procedure Combination (DPC) inpatient database, to obtain data from 2018 to 2019 [6,15,16]. This dataset comprises information obtained from over 1000 hospitals, including all 82 teaching hospitals, and covers half or more of inpatient cases throughout Japan. The available information included diagnosis and comorbidities upon hospital admission coded according to the International Classification of Disease and Related Health Problems, 10th Revision [14]. The datasets also included information on patients’ age, sex, body mass index (BMI), the Charlson comorbidity index [17], which was updated for risk adjustment [18], ADLs upon admission and discharge, and discharge status.

A total of 15,422,773 eligible cases were identified. The criteria for study inclusion included age ≥50 years, hospitalization ≥24 h, and patients undergoing any of the defined major surgeries. Major surgeries were previously defined as invasive operative procedures, including CABG, thoracic lobectomy, orthopedic surgery (hip or knee arthroplasty, laminectomy or spinal fusion, and surgery for hip fracture or dislocation), hepatopancreatobiliary surgery (hepatic lobectomy, cholecystectomy, and pancreatectomy), gastrointestinal resection (esophagectomy, gastrectomy, and colectomy), and urological surgery (cystectomy and nephrectomy) [15,19]. The exclusion criteria were as follows: second or later admissions; patients with incomplete information on BMI, the Barthel Index, and admission type (emergency admission or not); those who underwent multiple surgery types; and dependence on maintenance dialysis therapy. Dependence on hemodialysis or peritoneal dialysis was determined from the coding of patient care procedures (chronic maintenance hemodialysis with <4 h per session, ≥4 h but <5 h per session, ≥5 h per session or chronic maintenance hemodiafiltration or continuous peritoneal dialysis) [17].

Our study protocol was approved by the ethics committee of Tokyo Medical and Dental University. The need for informed consent was waived because the data were anonymized. The study was conducted in accordance with the ethical principles of the Declaration of Helsinki. The data used for the study analyses are available from the corresponding author S.M. upon reasonable request.

### Patient characteristics

We calculated the Barthel Index scores upon admission and discharge based on 10 functional abilities, including feeding, bathing, grooming, dressing, bowel control, bladder control, toileting, chair transfer, ambulation, and climbing stairs [20]. Barthel Index scores ranged from 0 to 100 at 5-point increments, with 0 indicating complete bed-ridden status and 100 indicating full independence of physical functions. Other patient information on admission included age, sex, BMI, dependence on urgent or maintenance kidney replacement therapy, updated Charlson comorbidity index, excluding renal disease [17,18], comorbidity, surgery type, admission type (emergency or elective admission), and fiscal year. We used the following age strata for the analyses: 50–59, 60–69, 70–79, and ≥80 years [21]. On the basis of previous studies, antihypertensive medications were classified into six groups, including TH, CCBs, RASis, MRAs, α-blockers, and β-blockers [22, 23]. Thiazide-like diuretics without a benzothiadiazide structure, such as indapamide and metolazone, were also categorized under TH. Only dihydropyridines were included as CCBs in this study. RASis included ACEis and ARBs. Notably, α/β-blockers were included under β-blockers. Loop diuretics were recognized when used both orally and intravenously.

### Outcomes

The primary outcome was the composite of overall inhospital death and decline in physical function, which was defined as a 5-point lower Barthel Index score [24] at discharge than at admission. Patients were followed up until discharge, transfer, or inhospital death.

### Data analyses

Data were presented as number (percentage) or median (interquartile range [IQR]). Logistic regression models were used to assess the association between antihypertensive medication and loop diuretics and the risk for mortality or functional decline, adjusting for potential confounding measurements, including age, sex, type of major surgeries, BMI, Barthel Index score upon admission, diabetes mellitus (DM), CVD, chronic kidney disease (CKD), emergency admission, and year of admission. We conducted subgroup analyses using logistic regression models to determine the association between antihypertensive class and outcomes according to major surgery type. All statistical analyses were performed using Stata version 15.0 (Stata Corp., College Station, TX, United States), with *P* < 0.05 indicating statistical significance.

## Results

### Patient characteristics

We identified 15,422,773 patients from a Japanese national inpatient database between 2018 and 2019 (Fig. S1). Subjects aged 50 years and older who underwent major operations during hospitalization lasting for at least 24 h were included (n = 1,283,540). Subsequently, those on their second or later admissions; those without complete information on BMI, Barthel Index score, or emergency admission; and those who underwent multiple surgeries or received maintenance dialysis were excluded. In total, 849,910 subjects were included in the analysis.

Table 1 summarizes the demographics and characteristics of the study cohort. Among the 849,910 patients, 473,327 (56%) were taking at least one class of antihypertensive medication. Antihypertensive users and nonusers had a median age (IQR) of 76 (69–82) years and 70 (63–78) years, female percentage of 49% and 50%, and median BMI of 23.4 (20.9–26.0) kg/m^2^ and 22.5 (20.2–24.9) kg/m^2^, respectively. A larger proportion of antihypertensive users than nonusers had a low Barthel Index score. The prevalence of comorbidities, including CVD, DM, and CKD, was higher among antihypertensive users than among nonusers. Patients were also stratified according to the type of major surgery that they underwent during hospitalization. Among the six types of major surgeries (CABG, thoracic lobectomy, orthopedic surgery, hepatopancreatobiliary surgery, gastrointestinal resection, and urological surgery), orthopedic surgery was the most common in both groups (45% and 37%, respectively), followed by gastrointestinal resection and esophagectomy (20% and 25%, respectively).

**Table 1.**
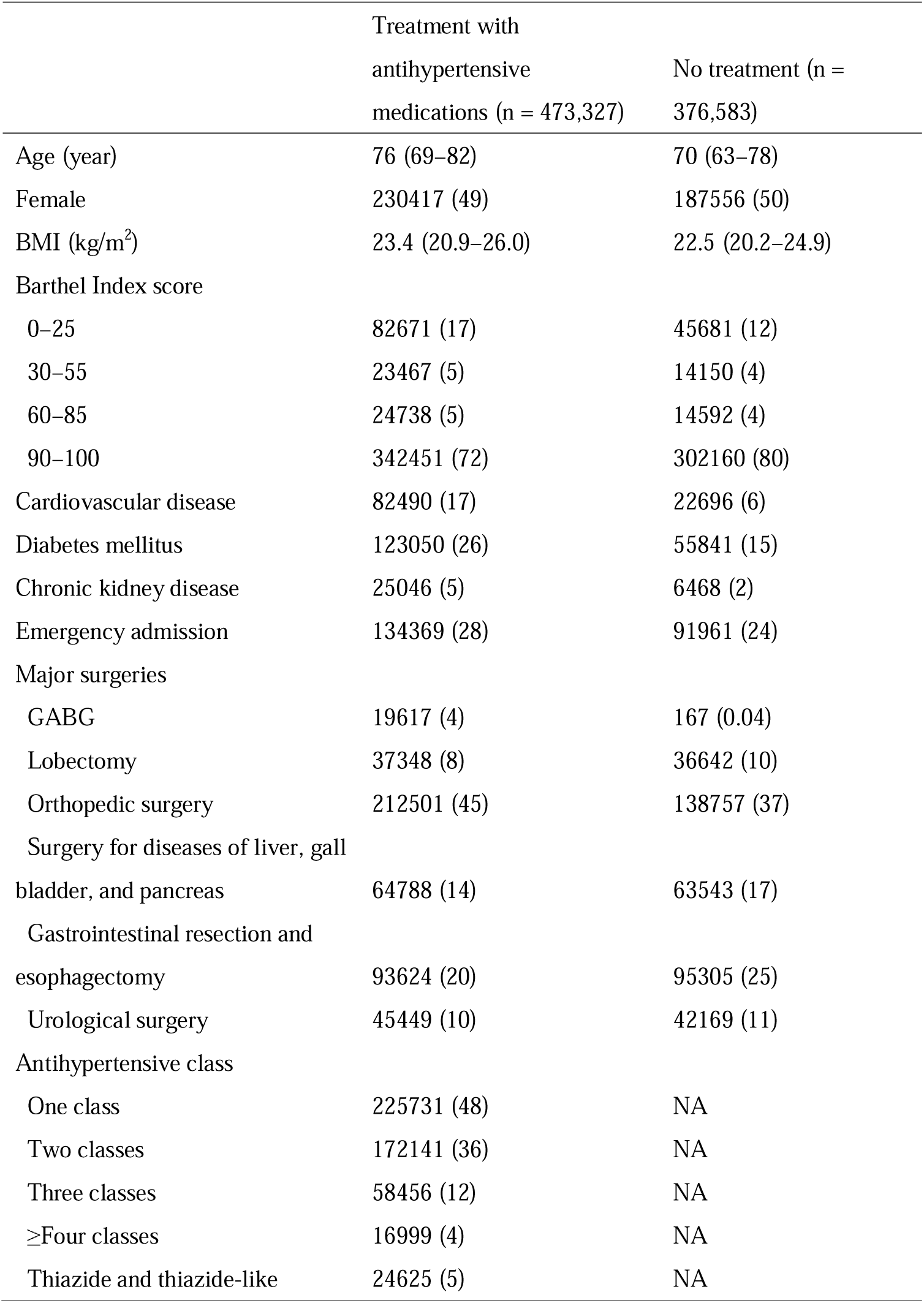
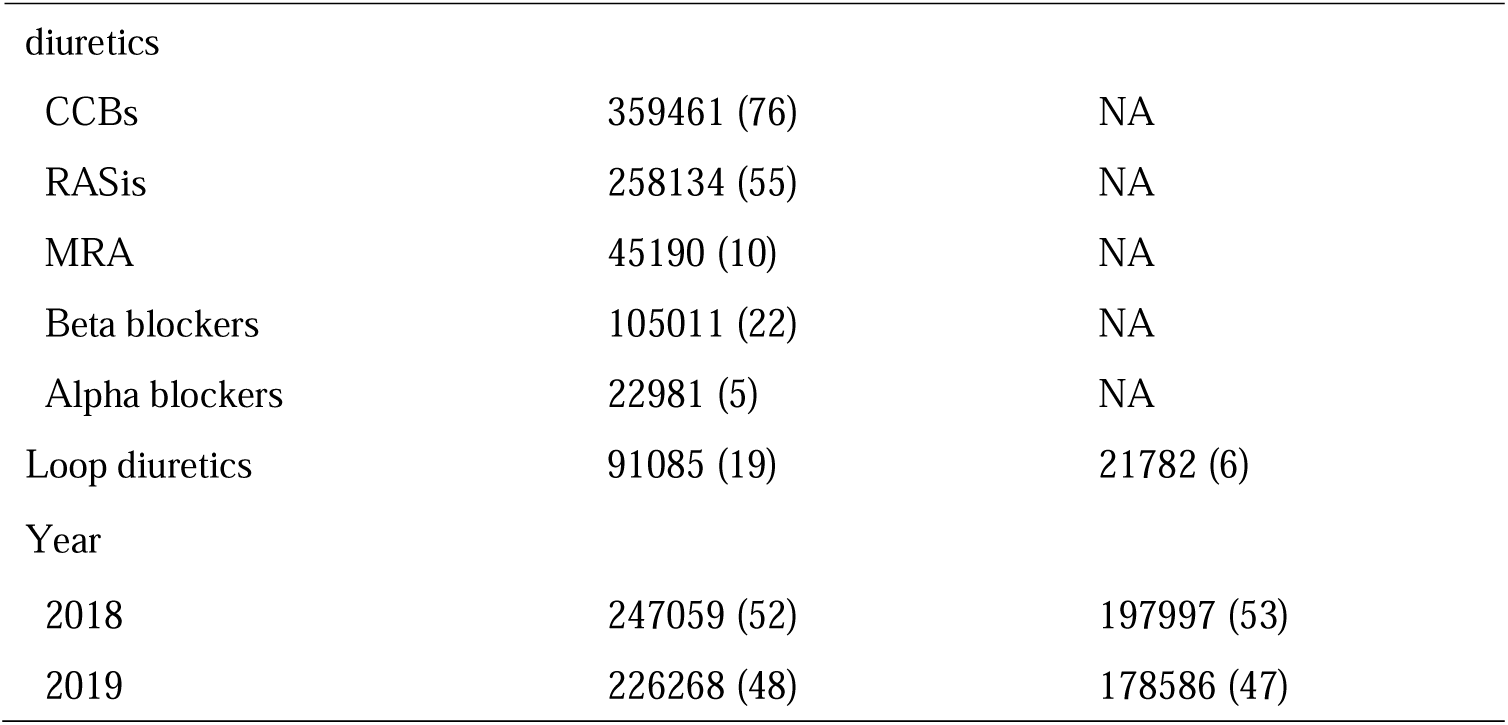
Characteristics of patients aged 50 years and older undergoing major. Data are presented as number (percentage) or median (interquartile range). BMI, body mass index; CABG, coronary artery bypass grafting; CCB, calcium channel blocker; MRA, mineral corticoid receptor antagonist; RASi, renin–angiotensin–aldosterone system inhibitor; NA, not applicable.

|  | Treatment with<br>antihypertensive<br>medications (n = 473,327) | No treatment (n =<br>376,583) |
| --- | --- | --- |
| Age (year) | 76 (69–82) | 70 (63–78) |
| Female | 230417 (49) | 187556 (50) |
| BMI (kg/m<sup>2</sup>) | 23.4 (20.9–26.0) | 22.5 (20.2–24.9) |
| Barthel Index score |  |  |
| 0–25 | 82671 (17) | 45681 (12) |
| 30–55 | 23467 (5) | 14150 (4) |
| 60–85 | 24738 (5) | 14592 (4) |
| 90–100 | 342451 (72) | 302160 (80) |
| Cardiovascular disease | 82490 (17) | 22696 (6) |
| Diabetes mellitus | 123050 (26) | 55841 (15) |
| Chronic kidney disease | 25046 (5) | 6468 (2) |
| Emergency admission | 134369 (28) | 91961 (24) |
| Major surgeries |  |  |
| GABG | 19617 (4) | 167 (0.04) |
| Lobectomy | 37348 (8) | 36642 (10) |
| Orthopedic surgery | 212501 (45) | 138757 (37) |
| Surgery for diseases of liver, gall<br>bladder, and pancreas | 64788 (14) | 63543 (17) |
| Gastrointestinal resection and<br>esophagectomy | 93624 (20) | 95305 (25) |
| Urological surgery | 45449 (10) | 42169 (11) |
| Antihypertensive class |  |  |
| One class | 225731 (48) | NA |
| Two classes | 172141 (36) | NA |
| Three classes | 58456 (12) | NA |
| ≥Four classes | 16999 (4) | NA |
| Thiazide and thiazide-like | 24625 (5) | NA |
| diuretics |  |  |
| CCBs | 359461 (76) | NA |
| RASis | 258134 (55) | NA |
| MRA | 45190 (10) | NA |
| Beta blockers | 105011 (22) | NA |
| Alpha blockers | 22981 (5) | NA |
| Loop diuretics | 91085 (19) | 21782 (6) |
| Year |  |  |
| 2018 | 247059 (52) | 197997 (53) |
| 2019 | 226268 (48) | 178586 (47) |

### Effects of the number of antihypertensive classes on postoperative outcomes

After a median follow-up of 17 (IQR, 10–27) days, all-cause inhospital deaths were observed in 5,777 patients (1.2%) out of 473,327 antihypertensive users and 2,657 patients (0.7%) out of 376,583 nonusers. Functional decline was observed in 22,550 (6.0%) nonusers and 42,930 (9.2%) users. Figure 1 displays the prevalence of all-cause inhospital death and functional decline after survival according to age category. The proportions of mortality and functional decline were higher among patients on more antihypertensive classes than among those on no or fewer antihypertensive classes. This indicates a significant association between resistance and antihypertensive treatment and the crude mortality rate and decline in ADLs (Figure 1)

**Figure 1.**
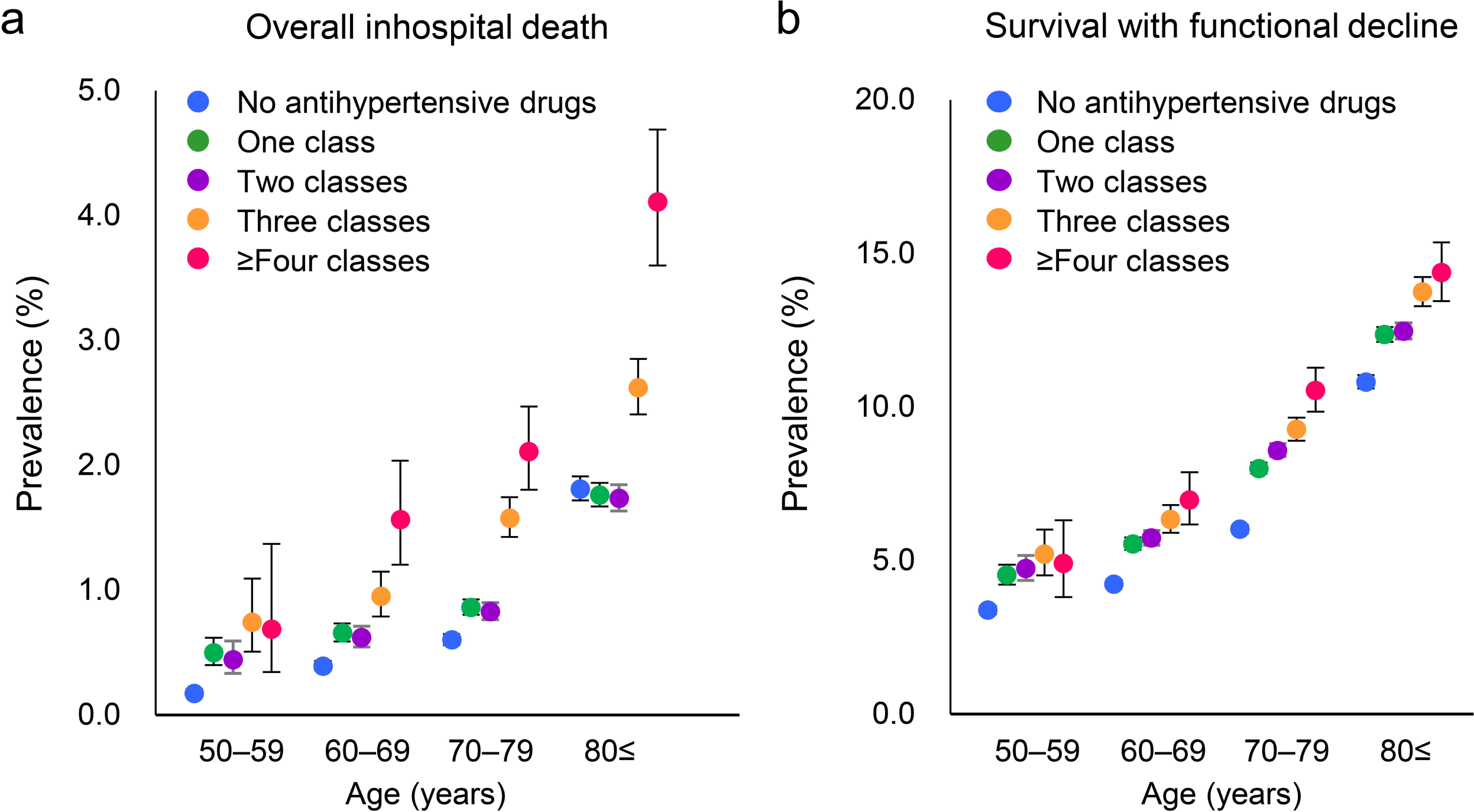
Antihypertensive classes and prevalence of postoperative death or functional decline after survival among patients undergoing major surgeries. **a, b.** prevalence of all-cause inhospital deaths (a) and functional decline (b) among nonusers and users of one, two, three, or four or more antihypertensive classes according to age category. Functional decline was defined as a ≥5-point decreased in the Barthel Index score between admission and discharge. Each spot represents a mean, whereas the solid lines represent the corresponding 95% CI. CI, confidence interval; n/N, number of events/number of participants.

Thereafter, multivariable logistic regression models were created to determine the aggregate risk for all-cause inhospital death and functional decline after survival. In the age- and sex-adjusted model, the odds ratios (ORs) for a composite outcome of mortality and functional decline were 1.20 (95% confidence interval [CI], 1.18–1.23), 1.23 (95% CI, 1.20–1.25), 1.42 (95% CI, 1.38–1.47), and 1.57 (95% CI, 1.49–1.65) for users receiving one, two, three, and four or more classes of antihypertensives when compared with nonusers (Table 2). Similarly, in Models 2 and 3, which adjusted for DM, CVD, CKD, emergency admission, and admission year, the use of more antihypertensive classes was associated with a higher risk for the composite outcome. After additionally adjusting for the use of loop diuretics (Model 4), the odds for the composite outcome were comparable between users receiving one, two, three, and four or more classes of antihypertensives. These findings indicate that congestion requiring the use of loop diuretics is a major confounding factor for the association between resistant hypertension and unfavorable postoperative outcomes.

**Table 2.** Number of antihypertensive classes and risk for mortality or functional decline after survival in patients undergoing major surgeries. Model 1: Adjusted for age, sex, and type of major surgeries (CABG, thoracic lobectomy, orthopedic surgery, hepatopancreatobiliary surgery, gastrointestinal resection, and urological surgery) Model 2: Model 1 plus BMI and Barthel Index score on admission. Model 3: Model 2 plus DM, CVD, CKD, emergency admission, and admission year. Model 4: Model 3 plus loop diuretics. CABG, coronary artery bypass grafting; BMI, body mass index; DM, diabetes mellitus; CVD, cardiovascular disease; CKD, chronic kidney disease; CI, confidence interval; OR, odds ratio.

|  | n/N | Model 1 | Model 2 | Model 3 | Model 4 |
| --- | --- | --- | --- | --- | --- |
| No antihypertensive drugs | 25207/376583 | Reference | Reference | Reference | Reference |
| One class | 22063/225731 | 1.20 (1.18–1.23) | 1.20 (1.17–1.22) | 1.17 (1.14–1.19) | 1.12 (1.10–1.14) |
| Two classes | 17558/172141 | 1.23 (1.20–1.25) | 1.20 (1.18–1.23) | 1.14 (1.12–1.17) | 1.14 (1.04–1.09) |
| Three classes | 6861/58456 | 1.42 (1.38–1.47) | 1.42 (1.38–1.46) | 1.30 (1.26–1.34) | 1.11 (1.08–1.15) |
| ≥Four classes | 2225/16999 | 1.57 (1.49–1.65) | 1.58 (1.50–1.67) | 1.43 (1.36–1.51) | 1.12 (1.07–1.19) |

### Combinations of antihypertensive classes and postoperative outcomes

We then investigated whether differences in postoperative outcomes following major surgeries were present according to different combinations of antihypertensive classes among the patients.

Among those who used only one of the six antihypertensive classes and loop diuretics, RASis showed the lowest OR for composite outcome, mortality, and functional decline (Table 3). With TH as the reference, RASi showed 0.77-, 0.58-, and 0.79-times lower odds for the composite outcome (95% CI, 0.63–0.93; *P* = 0.008), mortality (95% CI, 0.34–0.99; *P* = 0.045), and functional decline (95% CI, 0.64–0.96; *P* = 0.020), respectively, whereas β-blockers showed a 1.27-times higher odds for functional decline (95% CI, 1.01–1.60; *P* = 0.040; Table 3). Among those who used two antihypertensive classes, the combination of THs and RASi showed the lowest OR for each outcome. Compared with TH/CCB, TH and RASi had an OR of 0.68 for the composite outcome (95% CI, 0.60–0.77; *P* < 0.001; Table 3). Among those who used three or more classes of antihypertensive, combinations including CCB and RASi showed the lowest OR for each outcome. In comparison, using the combination of TH, CCB, and other as the reference, the combination of TH, CCB, and RASi showed odds of 0.72-, 0.43-, and 0.79-times lower for the composite outcome (95% CI, 0.49–0.82; *P* < 0.001), mortality (95% CI, 0.31–0.60; *P* < 0.001), and functional decline (95% CI,0.70–0.89; *P* < 0.001; Table 3), respectively. The use of RASis was significantly associated with a lower OR for postoperative outcomes regardless of the number of antihypertensive classes used for treatment. A comparison of those who did and did not use loop diuretics showed that loop diuretic use consistently resulted in significantly higher ORs for the composite outcome, mortality, and functional decline regardless of the number of other antihypertensive medications used (Table S1).

**Table 3.**
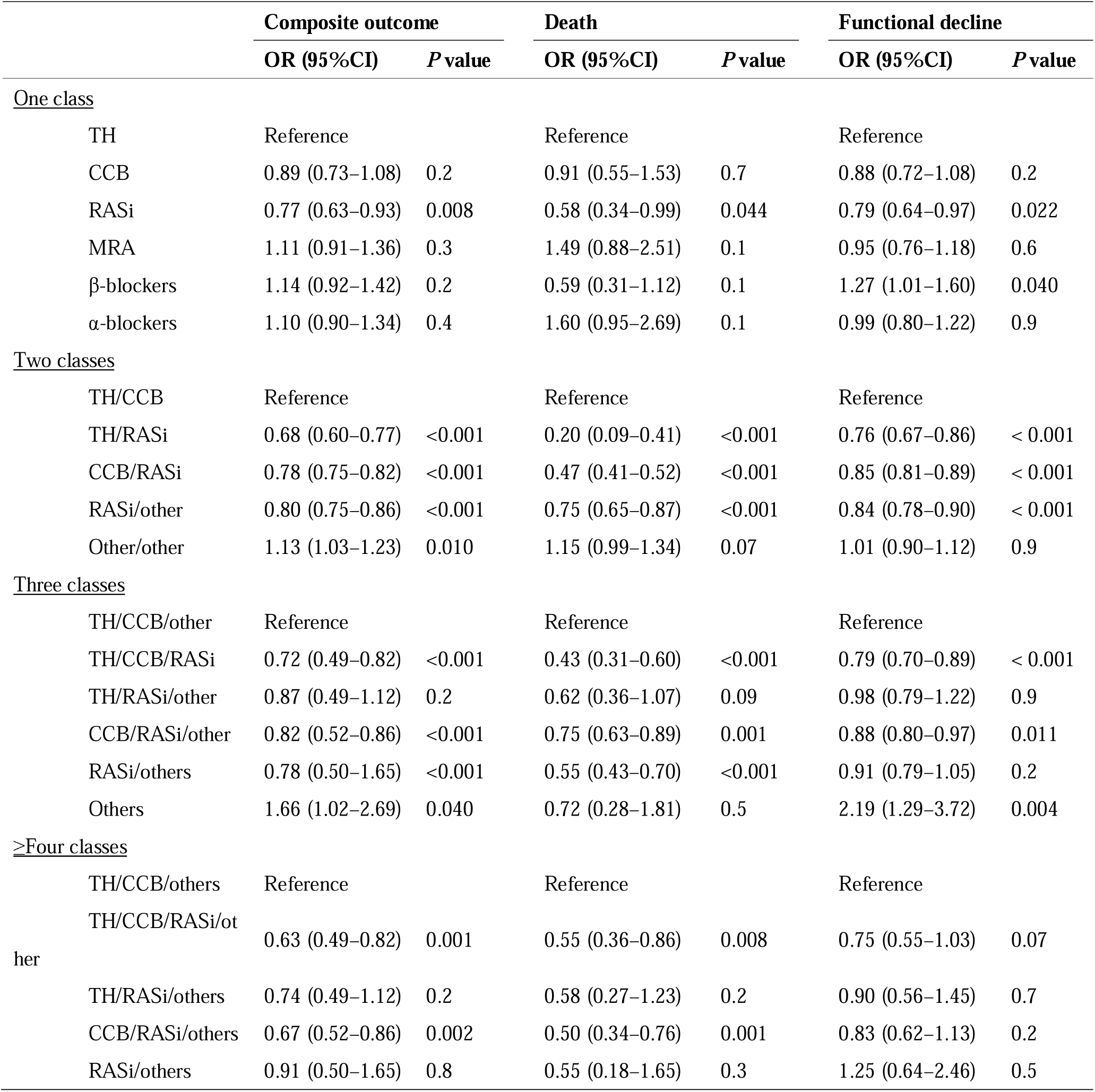
Combinations of antihypertensive classes and perioperative outcomes following major surgeries in Japanese adults. Multivariate logistic regression models adjusted for age, sex, type of major surgeries (CABG, thoracic lobectomy, orthopedic surgery, hepatopancreatobiliary surgery, gastrointestinal resection, and urological surgery), BMI, Barthel Index score on admission, DM, CVD, CKD, emergency admission, admission year, and loop diuretics. CABG, coronary artery bypass grafting; BMI, body mass index; DM, diabetes mellitus; CVD, cardiovascular disease; CKD, chronic kidney disease; CI, confidence interval; OR, odds ratio; CCB, calcium channel blocker; MRA, mineral corticoid receptor antagonist; RASi, renin–angiotensin–aldosterone system inhibitor; TH, thiazide/thiazide-like diuretics.

### Impact of each antihypertensive class on postoperative outcomes according to the type of major surgery

We performed a stratified analysis on the impact of each antihypertensive class on postoperative outcomes according to six types of major surgeries (CABG, thoracic lobectomy, orthopedic surgery, hepatopancreatobiliary surgery, gastrointestinal resection, and urological surgery). Notably, RASis were associated with lower ORs for the composite outcome of death and functional decline, all-cause inhospital death, and survival with functional decline among those who underwent five types of major surgeries except CABG (Figure 2). Only β-blocker use was associated with better outcomes among patients who underwent CABG (OR, 0.77; 95% CI, 0.67–0.89).

**Figure 2.**
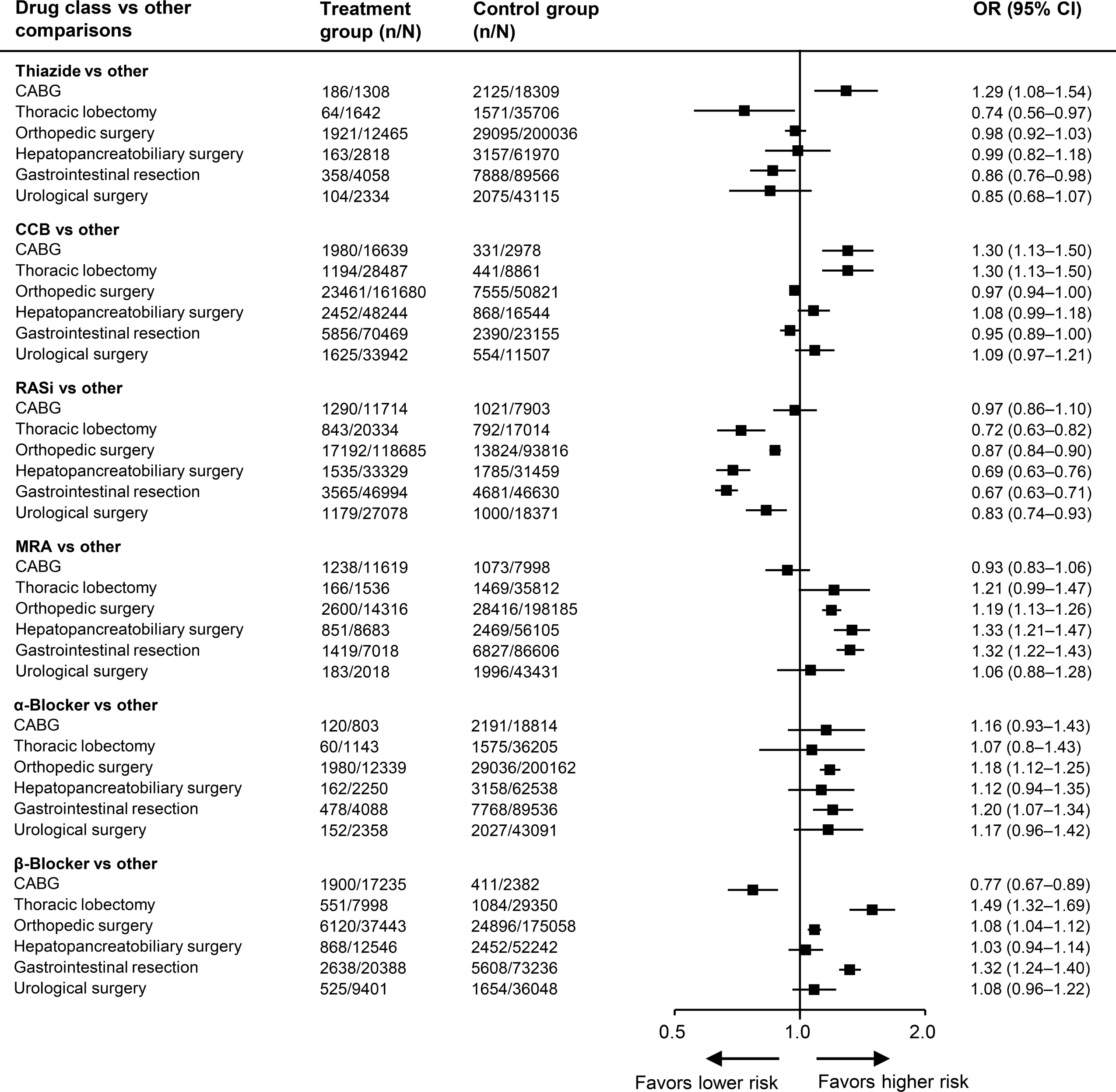
Association between surgery type and effects of antihypertensive classes on postoperative outcomes among patients with hypertension. Multivariable logistic regression models adjusted for age, sex, number of antihypertensive classes, BMI, Barthel Index score on admission, DM, CVD, CKD, emergency admission, year, and loop diuretics according to surgery type. The type of major surgeries included CABG, thoracic lobectomy, orthopedic surgery, hepatopancreatobiliary surgery, gastrointestinal resection, and urological surgery. CABG, coronary artery bypass grafting; BMI, body mass index; DM, diabetes mellitus; CVD, cardiovascular disease; CKD, chronic kidney disease; CI, confidence interval; OR, odds ratio; CCB, calcium channel blocker; MRA, mineral corticoid receptor antagonist; RASi, renin–angiotensin–aldosterone system inhibitor.

## Discussion

The current large-scale epidemiologic cohort study using the Japanese administrative claims database clarified the effects of antihypertensive drugs on postoperative mortality and functional decline in patients aged 50 and older. First, we found that an increase in the number of antihypertensive classes, which indicated treatment-resistant hypertension, was associated with a higher risk of mortality and loss of physical function, partly attributed to loop diuretic use needed for congestion. Second, we determined combinations of antihypertensives that potentially improve the outcomes of antihypertensive users undergoing major surgeries. The favorable regimens included RASis independent of the number of antihypertensive classes used. Third, among patients who underwent any type of major surgery, excluding CABG, those on RASis were at a lower risk of death and functional decline than those not treated with other antihypertensive classes.

The primary finding of the current study was that patients who received RAS inhibition had a lower risk for the composite outcome of all-cause inhospital death and functional decline after surviving major surgeries than those without RAS inhibition, independent of the number of antihypertensive classes used. RAS inhibition in patients treated with ACEi and ARB is known to help decrease skeletal muscle loss [7–9]. A plausible explanation for its underlying pathological mechanism is the suppression of angiotensin II-induced impairment in protein synthesis, acceleration of protein breakdown, and loss of appetite. Angiotensin II increases the levels of cytokines and systemic hormones, including reactive oxygen species, insulin-like growth factor 1, tumor necrosis factor alpha, interleukin-6, and transforming growth factor beta [7–9, 24–28]. The current study showed that among patients who used a single class of antihypertensive, those who used RASi had lower odds for the composite outcome of mortality and functional decline than those who used TH. Among those who received two classes of antihypertensives, those who received a combination of TH/RASi had an OR of 0.68 for the composite outcome compared with those on TH/CCB. The favorable effects of RASis on mortality and functional decline were also observed among those who used three or more antihypertensive classes. Our data clearly showed that RAS interfered with functional decline, although pathological analysis at the molecular level was not conducted.

Our findings showed that β-blocker users had a 1.27-fold higher OR for functional decline than TH users, which is consistent with previous findings on the association between β-blocker and functional decline in patients with end-stage kidney disease (ESKD) [29], CVD, and heart failure [30, 31]. In our previous study on a cohort with ESKD, β-blocker users had 2.5-fold higher odds of annual skeletal muscle mass loss, whereas the use of RASi in combination with other medications minimized the adverse effects [29]. Notably, only the use of β-blockers was markedly associated with a lower risk for postoperative composite outcome in patients who underwent CABG. Therefore, the current study reaffirms the positive effects of RASi on skeletal muscle mass in acute care settings among patients undergoing major surgeries [29].

Nonetheless, further investigations are needed to uncover the mechanism underlying the association between RAS inhibition and favorable outcomes after major surgeries, excluding CABG.

Our findings showed that outcomes following major surgeries could be further improved by combining empirically antihypertensive drugs prescribed. Frailty, often defined as unintentional weight loss, has been shown to be associated with mortality among inhospital patients [32], as well as postdischarge patients or outpatients [33–35]. Newly acquired functional decline among older persons owing to hospital admissions is especially serious given the already high prevalence of frailty among community-dwelling people in the aging Japanese society [36] and its association with higher mortality and economic burden. Our findings showed that 9.2% of antihypertensive drug users and 6.0% of nonusers experienced a decrease in their Barthel Index score during hospitalization, a figure even higher than the 7.4% overall pooled prevalence of frailty among Japanese community-dwelling people aged 65 and older reported in a previous review [36]. This suggests that the probability of functional decline is primarily higher among inpatients and that patients with hypertension are at substantially higher risk. Thus, combinations of antihypertensive drugs, including RASis, may improve the clinical outcomes of patients with hypertension by minimizing functional decline through internal treatment during the acute phase of hospitalization.

A substantial proportion of patients worldwide remain resistant to hypertension treatment and are usually treated with more antihypertensive medication classes.

However, the optimal combinations of antihypertensives are yet to be fully understood. As expected, our findings showed that the number of antihypertensive classes was positively correlated with the risk of mortality and functional decline. Indeed, we found that patients on multiple antihypertensive medications were at a greater risk for the composite outcome, even after adjusting for potential confounders. Notably, after adjusting for the use of loop diuretics, the odds for the risk of the composite outcome were similar regardless of the number of antihypertensive classes used, suggesting that loop diuretic usage was a powerful confounding factor associated with mortality and functional decline. Loop diuretics are indispensable in addressing fluid retention among patients with renal and heart failure. One possible side effect of loop diuretics is sarcopenia, as discussed in previous studies [10–14]. Na+-K+-2Cl-cotransporter 1, which is highly expressed in mammalian skeletal muscle and inhibited by furosemide, might help prevent muscle loss based on its background pharmacological mechanism [10]; however, the pathological pathway has yet to be not fully elucidated.

One strength of the current study is that we used well-validated Japanese administrative data representing a sample of the general population. The validity of the diagnoses and procedural records in the Japanese DPC database was corroborated in a previous study [37]. However, several limitations need to be acknowledged. First, this study was exclusively conducted on a single race or ethnicity. Evidence suggests that Asian populations have a higher prevalence of uncontrolled hypertension [38]. Hence, our findings may not be generalizable to other ethnicities. Second, the use of more than one antihypertensive drug within the same antihypertensive class was under-represented in this study. Third, given the nature of the datasets used herein, we were unable to determine changes in blood pressure and antihypertensive drug use during hospitalization potentially caused by poor control, complications, or side effects. The number of antihypertensive classes may have been overestimated. Finally, the database lacked biochemical data and longitudinal outcomes after discharge. Our study focused only on the effects of antihypertensive medication on postoperative mortality and functional decline during the acute phase of hospitalization following major surgeries. A previous multicenter retrospective cohort study showed that the postoperative initiation of RASis even at discharge was associated with reduced long-term mortality rate in patients undergoing cardiac surgery [39]. Randomized controlled trials are needed to determine whether perioperative RASi initiation could improve outcomes following cardiac and non-cardiac surgeries.

In conclusion, the current study highlights the effects of antihypertensive classes and their combinations on the risk of postoperative mortality and functional decline. Our findings showed that RASis usage had a positive prognostic impact on postoperative inhospital death and functional decline regardless of the number of antihypertensive medication classes used. Thus, RASi use may mitigate negative postoperative outcomes after major noncardiac surgeries.

## Data Availability

All data produced in the present study are available upon reasonable request to the authors.

## Acknowledgments

We would like to thank all study participants.

## Nonstandard Abbreviations and Acronyms

ACEi: angiotensin-converting enzyme inhibitor
AKI: acute kidney injury
ADL: activities of daily living
AHT: acute hypertension
ARB: angiotensin II receptor blocker
BMI: body mass index
CABG: coronary artery bypass grafting
CCB: calcium channel blocker
CI: confidence intervals
CKD: chronic kidney disease
CVD: cardiovascular disease
DM: diabetes mellitus
DPC: diagnosis procedure combination
ESKD: end-stage kidney disease
ICD-10: International Classification of Disease and Related Health Problems, 10th Revision
IQR: interquartile range
MRA: mineral corticoid receptor antagonist
OR: odds ratio
RAS: renin–angiotensin system
RASi: renin–angiotensin–aldosterone system inhibitor
ROS: reactive oxygen species
TH: thiazide/thiazide-like diuretics

