## Supplementary material for "Association between antihypertensive combinations and postoperative mortality and functional decline: a nationwide survey of Japanese adults undergoing major surgeries": Figure S1 and Table S1

**Figure S1. Patient flowchart.**

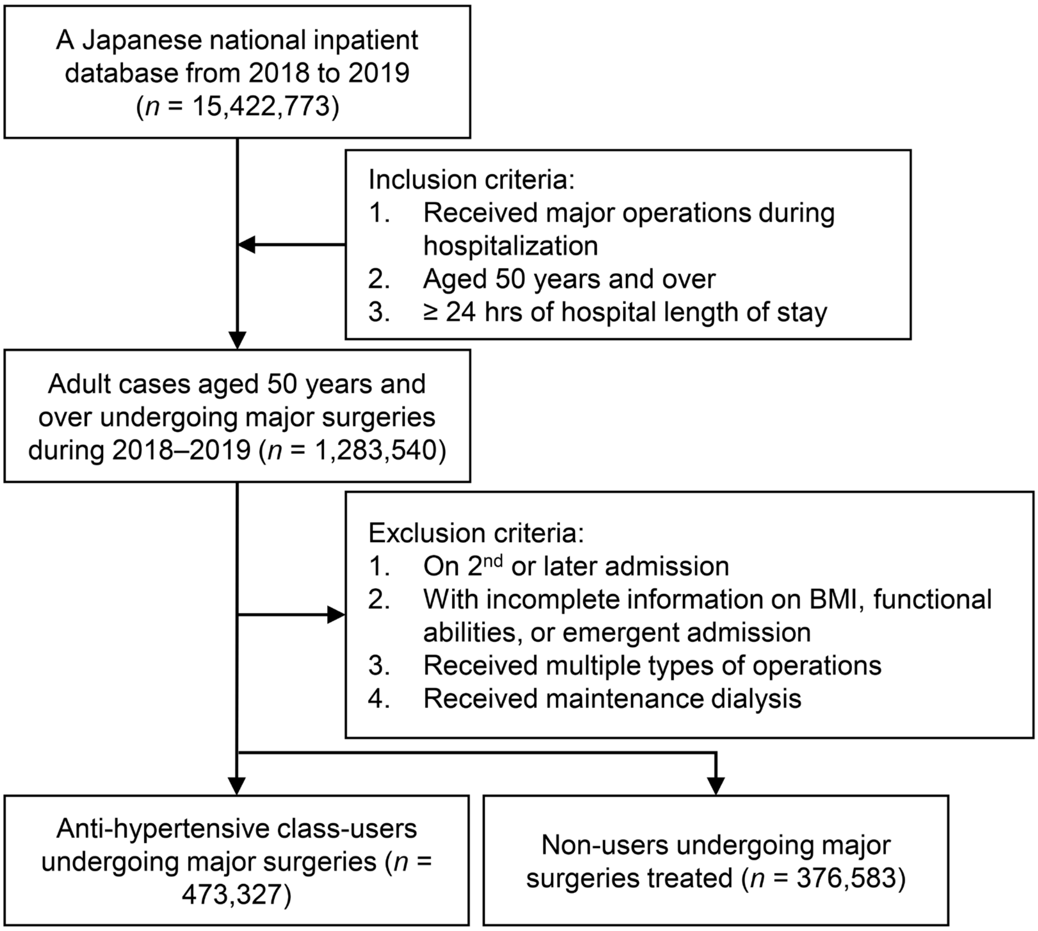

**Table S1. Combinations of antihypertensive classes and perioperative outcomes following major surgeries in Japanese adults.**

| | Composite outcome | | Death | | Functional decline | |
|---|---|---|---|---|---|---|
| | **OR (95%CI)** | ***P* value** | **OR (95%CI)** | ***P* value** | **OR (95%CI)** | |
| One class | | | | | | |
| Loop diuretic user vs. nonuser | 2.06 (1.98–2.15) | <0.001 | 5.68 (5.17–6.23) | < 0.001 | 1.56 (1.49–1.64) | < 0.001 |
| Two classes | | | | | | |
| Loop diuretic user vs. nonuser | 1.90 (1.82–1.98) | <0.001 | 5.60 (5.00–6.29) | <0.001 | 1.51 (1.44–1.59) | < 0.001 |
| Three classes | | | | | | |
| Loop diuretic user vs. nonuser | 1.77 (1.66–1.88) | <0.001 | 6.13 (5.16–7.28) | <0.001 | 1.35 (1.26–1.45) | < 0.001 |
| ≥Four classes | | | | | | |
| Loop diuretic user vs. nonuser | 1.84 (1.63–2.07) | <0.001 | 6.55 (4.58–9.36) | <0.001 | 1.42 (1.24–1.63) | <0.001 |

Multivariate logistic regression models adjusted for age, sex, type of major surgeries (CABG, thoracic lobectomy, orthopedic surgery, hepatopancreatobiliary surgery, gastrointestinal resection, and urological surgery), BMI, Barthel Index score on admission, DM, CVD, CKD, emergency admission, and admission year, and antihypertensive combinations. CABG, coronary artery bypass grafting; BMI, body mass index; DM, diabetes mellitus; CVD, cardiovascular disease; CKD, chronic kidney disease; CI, confidence interval; OR, odds ratio.
